# DYCAST early warning system for dengue virus risk areas, Ribeirão Preto, Brazil

**DOI:** 10.64898/2026.11.18.26368416

**Authors:** Ryan M. Carney

## Abstract

Dengue is currently the fastest-spreading vector-borne disease, with an estimated 100-400 million infections per year and nearly 6 billion people at risk – more than half the world’s population. However, there is no specific antiviral treatment, thus the primary method for preventing infection is through targeted control of the principal mosquito vector, *Aedes aegypti*. The objective of this retrospective study is to evaluate the efficacy of forecasting dengue risk using human cases and an adaptation of DYCAST: a biologically based, spatiotemporal clustering model that proved operationally successful as a West Nile virus early warning system. Results demonstrate that DYCAST provided accurate and early detection of high-risk areas for dengue in Ribeirão Preto, Brazil, including a cryptic interepidemic focus of transmission preceding a severe epidemic. Using suspected cases, DYCAST predicted 90.3% (4,237/4,690) of confirmed cases, with a mean lead time of 66.3 days prior to illness onset. High-risk cells had >10× higher relative risk of containing a confirmed case, and sensitivity and specificity were 83.8% and 72.3%. These findings suggest that DYCAST could be utilized in real time to prospectively identify areas at high risk of dengue, for rapid and precise targeting of mosquito mitigation and public education. Implementation using free, open-source software offers a scalable technological strategy for the prevention and control of dengue around the world.

This article is part of the theme issue ‘The Dengue Prevention Toolbox: Integrating Biological, Social and Technological Strategies for Sustainable Control’.

## 1. Introduction

Dengue, caused by the mosquito-borne dengue virus (DENV; genus *Orthoflavivirus*, family *Flaviviridae*), is currently the fastest-spreading vector-borne disease. Each year there are an estimated 100-400 million infections, and nearly 6 billion people – more than half of the world’s population – are at risk of infection (*1*). Exacerbated by growing trends of urbanization and environmental perturbation, this tremendous global disease burden falls almost exclusively on developing countries, which generally lack the resources and infrastructure for sufficient surveillance and control of the virus. Yet this is increasingly a problem for developed countries as well, as exemplified by the continental U.S. currently (2026) experiencing its largest dengue epidemic in nearly a century (*2*).

The World Health Organization’s (WHO) 1997 classification system included undifferentiated fever, dengue fever, and dengue hemorrhagic fever (DHF), including dengue shock syndrome (DSS) (*3*). In 2009, the WHO revised this system to guide triage and clinical management, distinguishing dengue without warning signs, dengue with warning signs, and severe dengue (*3*). An estimated 60–80% of DENV infections are asymptomatic or subclinical, while approximately 2–5% of symptomatic cases progress to severe dengue (*4*). Untreated severe dengue has a case-fatality rate of ≥10%, although appropriate clinical management, including intravenous rehydration, can reduce this to <1% (*3*,*5*). Secondary infection with a different serotype (DENV-1, -2, -3, and -4) increases the risk of severe dengue, in part due to antibody-dependent enhancement (ADE; *4*). The potential for ADE also complicates vaccine development and deployment, and there are currently no specific antiviral treatments available for dengue (*4*). Thus, control of the principal mosquito vector, *Aedes aegypti*, is central to preventing human morbidity and mortality. There is an acute need for an early warning system to target such prevention and control efforts, and in particular, to identify areas where the virus is maintained undetected (cryptically) during interepidemic periods (*6*).

From 1990-1991, the first dengue epidemic occurred in Ribeirão Preto (652 km²; population: 400K (1991), 560K (2006), 732K (2025); ibge.gov.br), a northern city in the State of São Paulo, Brazil. During this outbreak there were ∼8,900 reported cases (2,305 and 242 confirmed in 1990 and 1991; *7*), and a survey conducted in 1992 detected 5.4% seroprevalence for DENV-1 IgG antibodies among participating residents (*8*). DENV-2 and -3 were detected in 1998 and 2001, respectively, with the latter triggering a large epidemic during that same year (Benedito Antônio Lopes da Fonseca, University of São Paulo, pers. comm.). Since then, Ribeirão Preto has experienced epidemics of generally increasing magnitude, with peaks of confirmed cases in 2006 (6,051; Figure 1; Supplementary Table S1), 2010 (29,637), 2011 (23,384), 2016 (35,043), and 2024 (44,630; the highest to date) (*9*). Vector control efforts are initiated in the city once the monthly incidence rate exceeds the average rate from the previous 10 non-epidemic years (Claudio Souza de Paula, Secretaria Municipal de Saúde, pers. comm.). However, this practice only provides a temporal threshold for epidemic control, and ignores the spatial aspects of dengue transmission. Complicating surveillance and control efforts is the fact that the health departments and vector control agency had relied exclusively on paper-based mapping of cases and vector data (Supplementary Figure S1).

**Figure 1.**
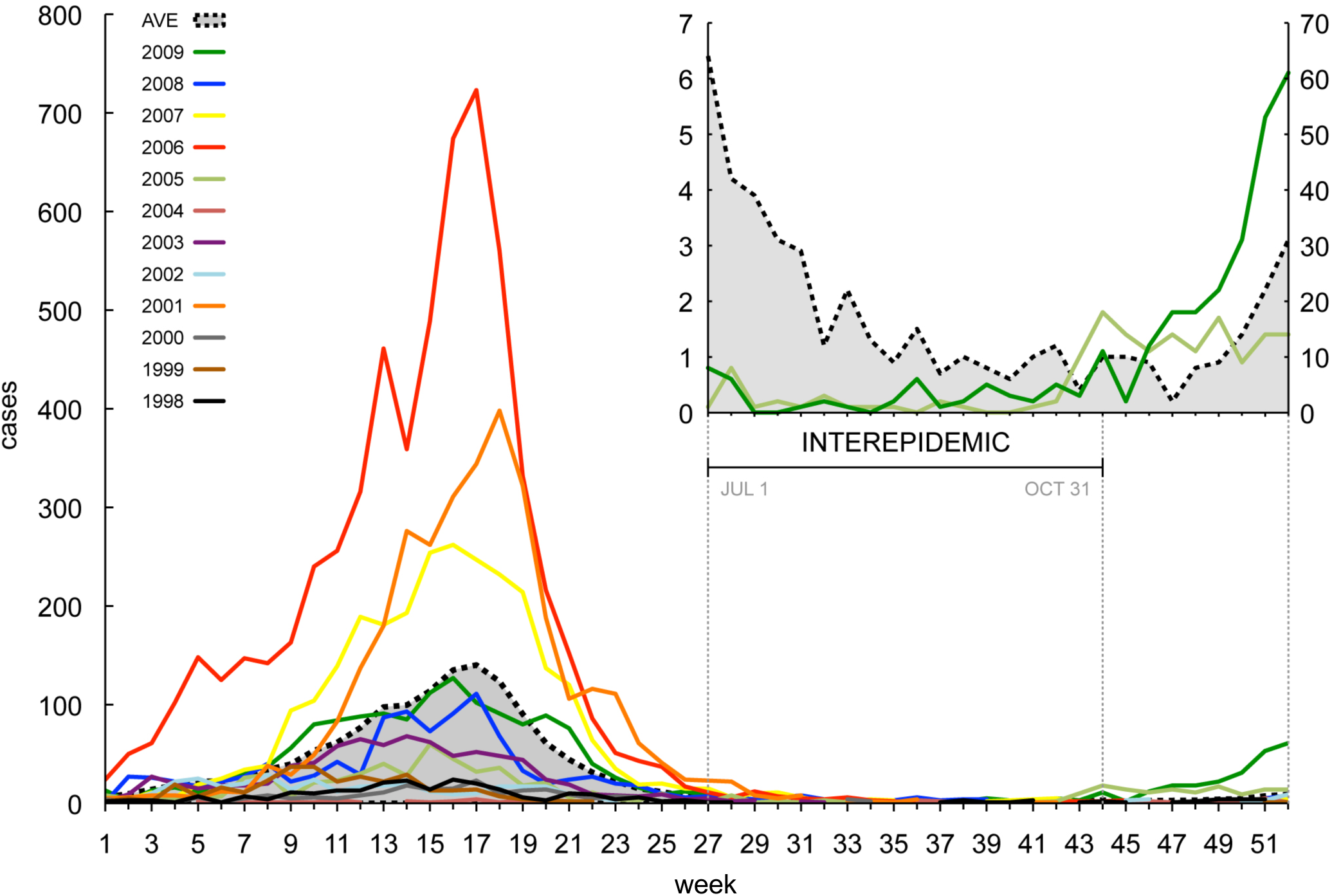
Dengue cases in Ribeirão Preto per week, by year. Average is the mean case incidence over the 12 years shown; average in the inset does not include 2005 and 2009 data, however. Inset x-axis has the same scale, Jul 1–Dec 31; inset y-axes have the same units (cases).

The objectives of this study were threefold. First, this study aimed to create a comprehensive GIS that could be utilized by local agencies in Ribeirão Preto for the surveillance and control of dengue, and in particular, a means to geocode the addresses of dengue cases for mapping purposes. Second, these spatial datasets would be used to implement a dengue risk model, which would be evaluated for use as a prospective, early warning system in Ribeirão Preto. The third objective was to use this model to detect cryptic transmission foci responsible for interepidemic maintenance of the virus during the dry season, which would be valuable for targeting mosquito control efforts to interrupt transmission prior to reemergence of epidemic amplification.

The majority of research on DENV transmission has focused on the epidemic periods of the disease, ignoring interepidemic transmission that nonetheless contributes considerable disease burden through productivity loss and absenteeism (*1*). What is known is that this phenomenon appears to be driven more by intrinsic population dynamics of the vector than extrinsic factors such as climate (*6*), suggesting that modeling efforts should concentrate more on intra-seasonal instead of inter-seasonal factors. However, in addition to numerous methodological shortcomings of using immature mosquito indices to predict risk of human infection (*10*), the vector surveillance data collected in Ribeirão Preto is prohibitive for using existing dengue transmission models (*11,12*) due to the entomological stage sampled (larval instead of pupal) and spatial scale at which data is aggregated (data not shown). Other researchers have used various spatial and spatiotemporal methods to characterize distributions of human cases (*13–16*), however all of these models have been only descriptive in nature and do not provide any statistical means for assessing future risk of dengue transmission.

However, previous studies have shown the Dynamic Continuous-Area Space-Time (DYCAST) model to be successful as an early warning system for the related mosquito-borne orthoflavivirus, West Nile virus (WNV). DYCAST identifies areas at high risk of WNV transmission to humans via a spatiotemporal cluster analysis of dead bird reports. Specifically, it uses a localized Knox test and Monte Carlo simulation to model viral amplification based on biological parameters of *Culex* spp. mosquito vectors and avian hosts. This model was first implemented in New York City in 2001, Chicago in 2002 (retrospectively), and California starting in 2005 as an early warning system (*17–19*). In California that year, DYCAST was used to assist mosquito larviciding and adulticiding efforts (*20*), including emergency aerial mosquito control conducted during an unprecedented epidemic, which has been shown to have reduced human morbidity and potentially mortality from WNV infection (*21*). Starting in 2006, DYCAST was implemented statewide as an official component of the California Mosquito-Borne Virus Surveillance and Response Plan (*19*). Given DYCAST’s demonstrated success as an early warning system for predicting and preventing a related mosquito-borne disease, this model was selected for the present study and adapted for use as a dengue risk model.

## 2. Material and methods

### 2.1. Data

Excel (Microsoft Corporation, Redmond, WA, USA) databases containing records of all suspected dengue cases (n=53,256) in Ribeirão Preto from January 1, 1998 through December 31, 2009 were obtained from the Ribeirão Preto Municipal Health District’s Division of Epidemiological Surveillance (Divisão de Vigilância Epidemiológica, Secretaria Municipal de Saúde de Ribeirão Preto). Information included residential address, date of onset of illness, and clinical diagnosis of confirmed (n=17,440; Supplementary Table S1) and unconfirmed (n=35,816) dengue cases. These data were cleaned and integrated into a single Excel database, and the confirmed cases used to plot case incidence by week in order to assess seasonality of epidemic and interepidemic periods. Locations of all suspected cases were mapped by using ArcView 9.3.0 (Environmental Systems Research Institute, Inc.; ESRI, Redlands, CA, USA) and a composite address locator to geocode records based on street address, Código de Endereçamento Postal (CEP; zip code), and bairro (district) (Supplementary Figure S2a); this locator was built by using a line shapefile containing street reference data for Ribeirão Preto and a Brazilian-formatted locator index, both obtained from ESRI. One source of spatial inaccuracy in geocoding address data is the offset value used to specify distance of residences from the centerline of their respective street, which can vary greatly between rural and urban areas. Thus, an appropriate offset was derived using empirical data, which consisted of 11,909 suspected dengue cases in Ribeirão Preto, from approximately January 1, 2007 through May 31, 2008, that had been previously georeferenced to centroids (center) of residential lots by Companhia de Desenvolvimento Econômico de Ribeirão Preto (CODERP; Wilson Yong, unpub. data). The median distance between cases and their respective street centerline was then obtained (Supplementary Figure S2b); cases >100 m from a street centerline were excluded, as these represented addresses not included in the street reference data. The median offset value was used in all subsequent geocoding processes.

Selection of address locator matching parameters (spelling sensitivity, minimum candidate score, and minimum matching score) was made through a step-wise sensitivity analysis. This included comparing geocoding results from all suspected cases, using both individual address locators and 14 combinations of parameters with the composite locator. An optimal parameter set was selected by examining results for accuracy. Approximately half (n=7,500) of the unmatched records were then manually rematched; findings from this process were then used to automatically rematch the remainder. Pearson correlation was used to test whether the percentage of successfully matched addresses changed over time, which might indicate changes in data quality or the presence of addresses in areas not represented by street reference data.

### 2.2. Risk model

#### 2.2.1. Software

In previous studies, the DYCAST model was implemented by using expensive and specialized GIS software, Smallworld 3.2.1 (General Electric Company, Fairfield, CT, USA), and its proprietary Magik computer programming language (*17–19*). More recently, the model has been recreated by using the widely used computer programming language, Python (www.python.org), which provides for the implementation of the model using free, open source, and cross-platform (Windows and Mac operating systems) software (Alan McConchie, University of British Columbia, unpub. data). The present study implemented this open source version of the model on a 64-bit Windows Vista Business Edition platform (Microsoft Corp.). DYCAST scripts were modified in order to run the model using dengue-specific parameters and the spatial reference appropriate for the study region; scripts were executed using Python 2.6. psycopg 2.0.14 (www.initd.org/psycopg) provided porting to PostgreSQL 8.4.3 object-relational database management system (www.postgresql.org); a graphical user interface (GUI) was provided by pgAdminIII (Supplementary Figure S3). Support for geographic objects was added by using PostGIS 1.5.1 (www.postgis.org) and shp2pgsql graphical loader plugin. The model’s analysis grid, risk maps, and animations were created by using ArcView 9.3.0, along with its Spatial Analyst and Tracking Analyst extensions. While ArcView is commercial software, these operations could be accomplished in future implementations by using the free and open source GIS software, Geographic Resources Analysis Support System GIS (GRASS GIS; grass.osgeo.org).

#### 2.2.2. Procedure

A digital map of Ribeirão Preto was superimposed with a grid consisting of 71,824 1-hectare (0.01 km^2^) cells. A localized Knox test (*22–23*) was implemented from the centroid of each cell for which the number of dengue cases within the spatial and temporal domains met or exceeded the analysis threshold (see below). The radius of the spatial domain was based on a 600 m maximum flight range of *Ae. aegypti* (*24,25*), a distance which fell within estimates of 200 m (*26*) and 800 m (*27*) derived from suburban and urban field studies in Rio de Janeiro, Brazil using laboratory-raised mosquitoes. The 28-day temporal domain represented one complete human-vector-human infection cycle (Figure 2); this accounted for a 10-day extrinsic incubation period of DENV in *Ae. aegypti* (*28–30*) and two human infection cycles of 9 days each, which consist of a 6-day intrinsic incubation period (*31–32*) and a 3-day infectious symptomatic period (*32*). These bounds define the spatiotemporal domain, within which cases were analyzed for close pairing in space and time. Closeness in space was defined as 100 m, to encompass the mean flight range of *Ae. aegypti* (*26,24,33,34*). Closeness in time was defined as 4 days, and represented the length of the *Ae. aegypti* gonotrophic cycle based on field (*35,36*) and laboratory (*33,37*) findings at temperatures consistent with those in Ribeirão Preto (Supplementary Figure S4). Statistical significance of case pairing was assessed by using unconditional Monte Carlo simulations (*18,38*). P-values denote the probability that clustering in space and time is caused by random chance; high-risk was defined at α=0.1 in accordance with previous DYCAST models (*17–19*). This procedure was repeated at each centroid to create an interpolated surface of risk. Parameters used in the WNV and DENV models are shown in Table 1 for comparison purposes.

**Figure 2.**
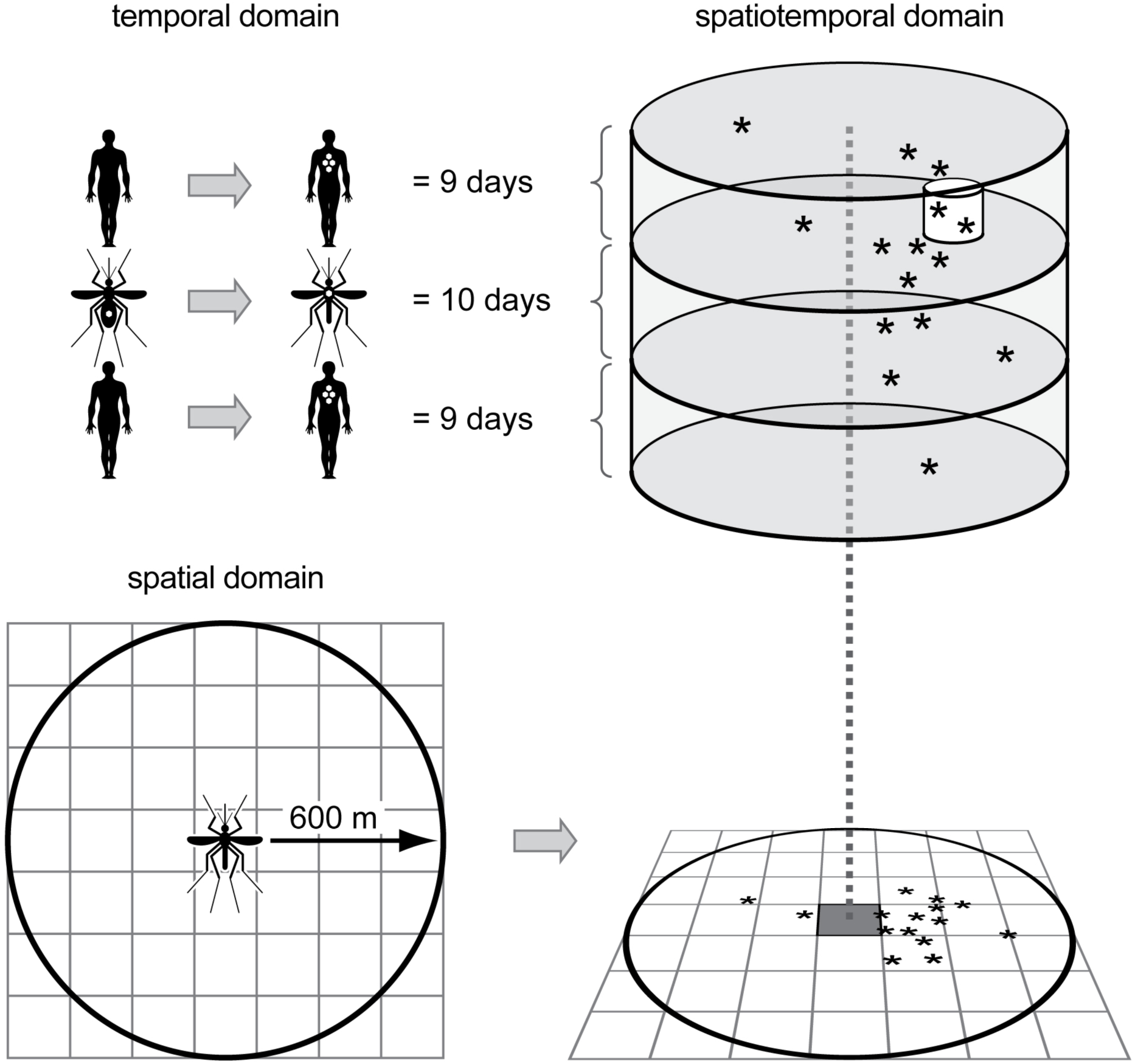
Schematic of the dengue virus DYCAST procedure, illustrating domains of localized Knox test implemented at the centroid of an individual 1-hectare grid cell (grid not to scale). The radius of the ≈113-hectare (1.13 km^2^) spatial domain is based on the maximum flight range of *Aedes aegypti*, and the 28-day temporal domain accounts for the extrinsic incubation period of DENV and two human infection cycles. These bounds define the spatiotemporal domain, within which cases (asterisks) were analyzed for close pairing in space (100 m) and time (4 days) (small white cylinder). Statistical significance of case pairing was assessed by using unconditional Monte Carlo simulations (α=0.1).

**Table 1.**

| Virus model | Spatial domain (m) | Spatial closeness (m) | Temporal domain (days) | Temporal closeness (days) | Threshold (number reports) |
|---|---|---|---|---|---|
| West Nile | 2,410 | 402 | 21 | 3 | 15 |
| dengue | 600 | 100 | 28 | 4 | 10,5 |

\*West Nile virus parameters taken from (*19*).

#### 2.2.3. Calibration

In order to determine an appropriate analysis threshold value, a sensitivity analysis was conducted for a date representative of an incipient epidemic (January 31, 2006). This included examinations of high-risk areas and p-value distributions generated by the DYCAST model at thresholds of 1 through 15. Optimal threshold(s) were subsequently used to run risk for the duration of the study period, for which year 2006 was selected as it represented the largest dengue epidemic in Ribeirão Preto in the dataset. The preceding interepidemic period in 2005 was also included in the study period, the exact date of which was determined by examination of case incidence to validate current temporal definitions. In Ribeirão Preto, the interepidemic period is officially defined as July 1 through October 31 of each year, approximately weeks 27 through 43 (Maria Luiza, Secretaria Municipal de Saúde de Ribeirão Preto, pers. comm.). Ecologically, the beginning of the subsequent epidemic period coincides with the rainy season which begins around November (de Paula, pers. comm).

#### 2.2.4. Evaluation

Results from the risk models were analyzed by using ArcMap, Excel:mac 2008 version 12.2.4, and StatPlus:mac LE 2009 version 5.8.0.0 (AnalystSoft, Inc., Vancouver, BC, Canada). Prediction was defined as the identification of a cell as high risk prior to or on the date of onset of illness of the earliest confirmed case located within that cell. If a cell was identified as high risk after the date of onset of illness, or the cell was never identified as high risk and a case occurred within it, it was designated false negative. Out of the total area analyzed, cells considered to be potentially at risk were those within 618.44 m of street reference data. This distance was based on the model’s 600 m spatial domain plus the 18.44 m geocoding side offset (see below), and represented the maximum distance from the street centerline that a cell could be identified as high-risk. This provided for a more conservative estimate of the true negative rate (and thus validity indices) by eliminating areas of the city where cases could not be mapped, and resulted in a study region of 25,487 cells, a total of 254.9 km^2^.

Results were used to calculate metrics of model efficacy, such as prediction rates and number of days prior to onset of illness that cases were predicted. Confusion matrices were also constructed to compare number of cells that contained dengue cases and number of cells identified as high risk by each model. These were used to generate measures of validity and association, including sensitivity, specificity, Youden’s J, positive (PPV) and negative predictive values (NPV), classification accuracy, relative risk (RR), and kappa statistic of chance-adjusted agreement (*39*). Because kappa is highly sensitive to both prevalence and classification bias, a prevalence-adjusted, bias-adjusted kappa statistic was used (PABAK; *40*). Results were also used to generate receiver operating characteristic (ROC) plots, and the area under the ROC curve (AUC) was estimated nonparametrically by using the Wilcoxon rank-sum statistic (*41*).

## 3. Results

### 3.1. Calibration

Results of weekly incidence plotted by year confirm the validity of the July 1 through October 31 temporal bounds of the interepidemic period; specifically, week 44 corresponds to increased incidence rates in 2005 and 2009 (Figure 1, inset), which preceded the unprecedented epidemics of 2006 and 2010 (9). Thus, July 1, 2005 through June 30, 2006 was selected as the duration for the model study period.

Geocoding offset analysis yielded a median distance of 18.44 m (60.50 feet) from residential lot centroid to street centerline; this value was used as the side offset value in the subsequent geocoding of dengue cases (Supplementary Figure S2a). Using the default parameters, geocoding using the composite locator provided a 1 percentage point increase in matched records compared to the CEP locator alone (Supplementary Table S2). Findings from the sensitivity analysis indicated that out of the 14 different combinations of matching parameters, 60, 10, and 23 was the optimal combination of spelling sensitivity, minimum candidate score, and minimum match score, respectively (Supplementary Table S2). Lowering the spelling sensitivity below 60 increased the number of inaccurate matches, due to a greater number of tied records (which are included in the total number of matches) that would otherwise be classified as unmatched. Subsequent manual examination and rematching of unmatched records indicated that the optimal parameter combination should be modified further to 60, 10, and 10. This rematching process also increased the total number of matched records from 72% to 73%. Results from the Pearson correlation test indicated no significant linear correlation between percentage of geocode matches and year (p=0.1174, R=-0.47644).

Results from the threshold sensitivity analysis indicated the occurrence of false positives and pair dependencies at low threshold numbers, which is an inherent statistical artifact of the Monte Carlo distributions. At a threshold of 2, for example, high-risk cells were predicted around every pair of cases (Figure 3a). However, this effect was eliminated at thresholds ≥10, which resulted in all high-risk cell p-values ≤ α of 0.1 (Figure 3b). Thus, a threshold of 10 was used for the initial risk model, D10. Additionally, in order to investigate the utility of using a lower threshold to increase sensitivity of the model during interepidemic periods, a threshold of 5 was used for a second risk model, D05. This threshold was based on an acceptable proportion of high-risk cell p-values ≤ α (88.1%; 998 of 1,133 cells), as well as qualitative examination of the risk maps, which indicated a sufficient balance of over- and under-prediction of risk at this threshold (Figure 3). A third model (D10’) was also run which used as its input all suspected cases of dengue, which included 4,496 unconfirmed cases in addition to the 4,690 confirmed cases used in the other models. The purpose of this iteration was to evaluate the effect that using raw syndromic surveillance data would have on the model’s efficacy.

**Figure 3.**
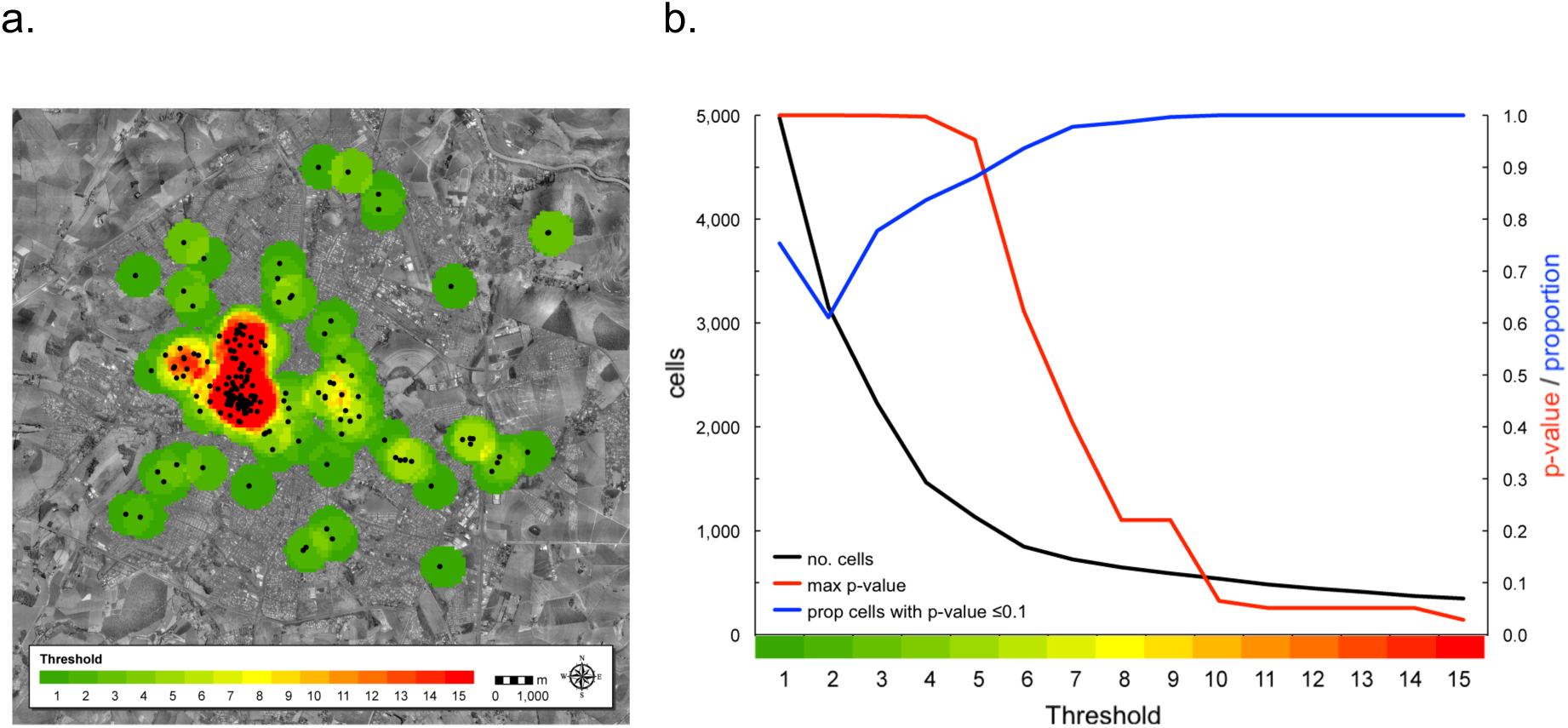
High-risk cells generated at analysis thresholds 1–15 for Jan 31, 2006, Ribeirão Preto, Brazil. a. Map illustrating dengue cases (black circles) with an onset of illness between Jan 3–Jan 31; this corresponds to the 28-day temporal domain of the model and thus includes all cases contributing to risk at that time. b. Graph illustrating number of high-risk cells, maximum p-value, and proportion of p-values ≤ α (0.1).

### 3.2. Evaluation

Models D10’, D05, and D10 first identified cells as high-risk on July 6, 14, and October 31; overall, cells were identified as high risk for a mean total of 92.3, 83.8, and 65.4 days, respectively (Tables 2, 3). During the study period (July 1, 2005 through June 30, 2006) there were 4,690 confirmed dengue cases with geocodable addresses, of which a maximum 90.3% (4,237/4,690) was predicted by the models at a mean of 66.3 days prior to onset of illness (D10’). Validity indices derived from confusion matrices (Table 2) yielded maximum sensitivity of 83.8% (D10’) and maximum specificity of 78.8% (D10); model D10’ exhibited the highest Youden’s J statistic, 0.561. Both measures of agreement, accuracy (0.777) and kappa (0.554), as well as PPV (24.2%), were highest in D10; NPV was highest in D10’ (97.8%). All three RR values differed significantly from unity, with a maximum of 10.569 (95% CI 9.498–11.760, p<0.0001) for D10’. ROC analysis yielded an AUC of 0.783 (Figure 4); given the similar results obtained by D05 and D10’, exclusion of the latter from the calculation yielded a virtually equivalent AUC of 0.780.

**Table 2.** Confusion matrices comparing number of cells that contained dengue case(s) and number of cells identified as high risk by models, 2005 Jul 1–2006 Jun 30, Ribeirão Preto*

|  |  | Model* |  |  |  |  |  |  |  |  |  |  |
|---|---|---|---|---|---|---|---|---|---|---|---|---|
|  |  | D10 |  |  |  | D05 |  |  |  | D10’† |  |  |
|  |  | Contained case |  |  |  | Contained case |  |  |  | Contained case |  |  |
|  |  | Yes | No | Total |  | Yes | No | Total |  | Yes | No | Total |
| High risk | Yes | 1,568 | 4,915 | 6,483 | Yes | 1,944 | 6,647 | 8,591 | Yes | 1,959 | 6,412 | 8,371 |
|  | No | 770 | 18,234 | 19,004 | No | 394 | 16,502 | 16,896 | No | 379 | 16,737 | 17,116 |
|  |  | 2,338 | 23,149 | 25,487 |  | 2,338 | 23,149 | 25,487 |  | 2,338 | 23,149 | 25,487 |

\* True positive (Yes/Yes) designates cell identified by DYCAST as high risk prior to or on the date of onset of illness of earliest case located within cell. If cell was identified as high risk after date of onset of illness, or cell was never identified as high risk and a case occurred within it, it was designated false negative (No/Yes). Number of cells that contained cases is less than the number of confirmed cases (4,690) due to 50.1% of cases occurring in cells containing other cases.
†D10 and D05 used confirmed dengue cases (n=4,690) and a threshold of 10 and 5, respectively; D10' used suspected dengue cases (n=9,186; both confirmed and non-confirmed cases) and a threshold of 10
‡Denotes results from model using a threshold of 10 and all suspected dengue cases (n=9,186) instead of only confirmed cases.

**Table 3.**
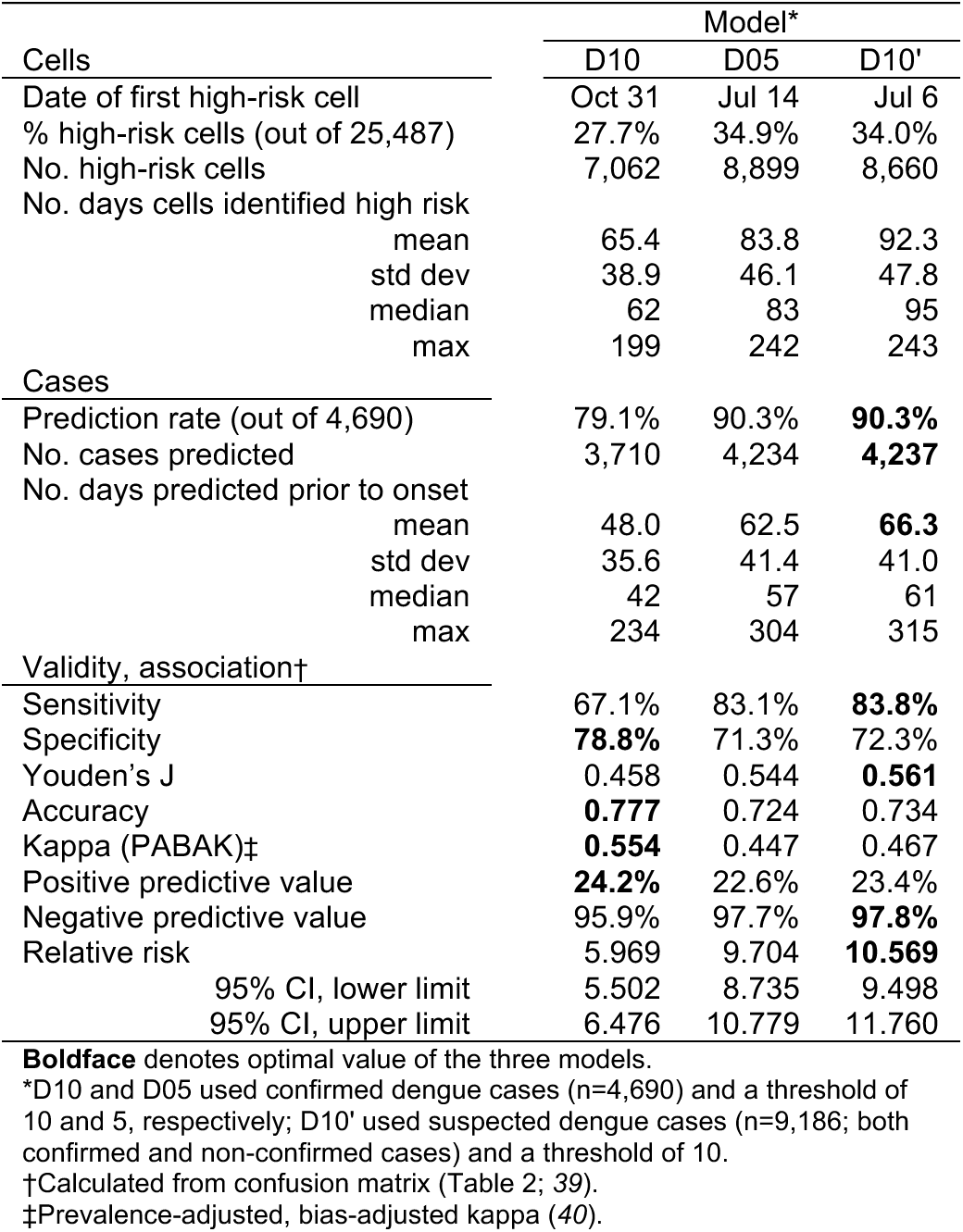
Model results, 2005 Jul 1–2006 Jun 30, Ribeirão Preto.

|  | Model* |  |  |
|---|---|---|---|
| Cells | D10 | D05 | D10' |
| Date of first high-risk cell | Oct 31 | Jul 14 | Jul 6 |
| % high-risk cells (out of 25,487) | 27.7% | 34.9% | 34.0% |
| No. high-risk cells | 7,062 | 8,899 | 8,660 |
| No. days cells identified high risk |  |  |  |
| mean | 65.4 | 83.8 | 92.3 |
| std dev | 38.9 | 46.1 | 47.8 |
| median | 62 | 83 | 95 |
| max | 199 | 242 | 243 |
| Cases |  |  |  |
| Prediction rate (out of 4,690) | 79.1% | 90.3% | **90.3%** |
| No. cases predicted | 3,710 | 4,234 | **4,237** |
| No. days predicted prior to onset |  |  |  |
| mean | 48.0 | 62.5 | **66.3** |
| std dev | 35.6 | 41.4 | 41.0 |
| median | 42 | 57 | 61 |
| max | 234 | 304 | 315 |
| Validity, association† |  |  |  |
| Sensitivity | 67.1% | 83.1% | **83.8%** |
| Specificity | **78.8%** | 71.3% | 72.3% |
| Youden’s J | 0.458 | 0.544 | **0.561** |
| Accuracy | **0.777** | 0.724 | 0.734 |
| Kappa (PABAK)‡ | **0.554** | 0.447 | 0.467 |
| Positive predictive value | **24.2%** | 22.6% | 23.4% |
| Negative predictive value | 95.9% | 97.7% | **97.8%** |
| Relative risk | 5.969 | 9.704 | **10.569** |
| 95% CI, lower limit | 5.502 | 8.735 | 9.498 |
| 95% CI, upper limit | 6.476 | 10.779 | 11.760 |

**Boldface** denotes optimal value of the three models.
\*D10 and D05 used confirmed dengue cases (n=4,690) and a threshold of 10 and 5, respectively; D10' used suspected dengue cases (n=9,186; both confirmed and non-confirmed cases) and a threshold of 10.
†Calculated from confusion matrix (Table 2; *39*).
‡Prevalence-adjusted, bias-adjusted kappa (*40*).

**Figure 4.**
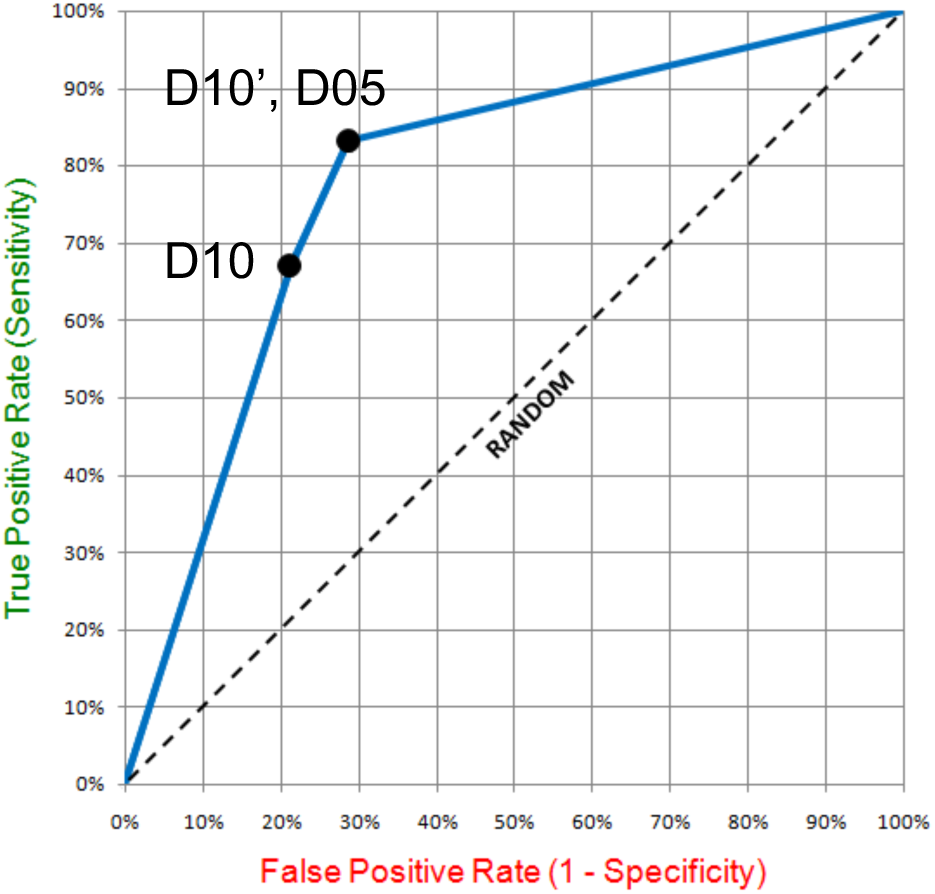
Receiver operating characteristic (ROC) plot for models D10, D05, and D10’. Area under the ROC curve (AUC) is equal to 0.783.

One interesting finding is that D10’ detected a cluster of cases in the western portion of the city at the beginning of the interepidemic period, identifying high-risk cells from July 6– August 8, 2005 (D05 also detected this cluster, but only identified risk between July 14–17) (Figure 5). What is important about this result is that this cluster was in the same location as the sole cluster of incipient risk identified in late October of that same year (Figure 6). Examination of the daily risk maps suggests that this October cluster was the initial focus of transmission for the 2006 epidemic, appearing to spill over into the surrounding areas (see Supplementary Video for an animation of D10’ risk maps during entire study period).

**Figure 5.**
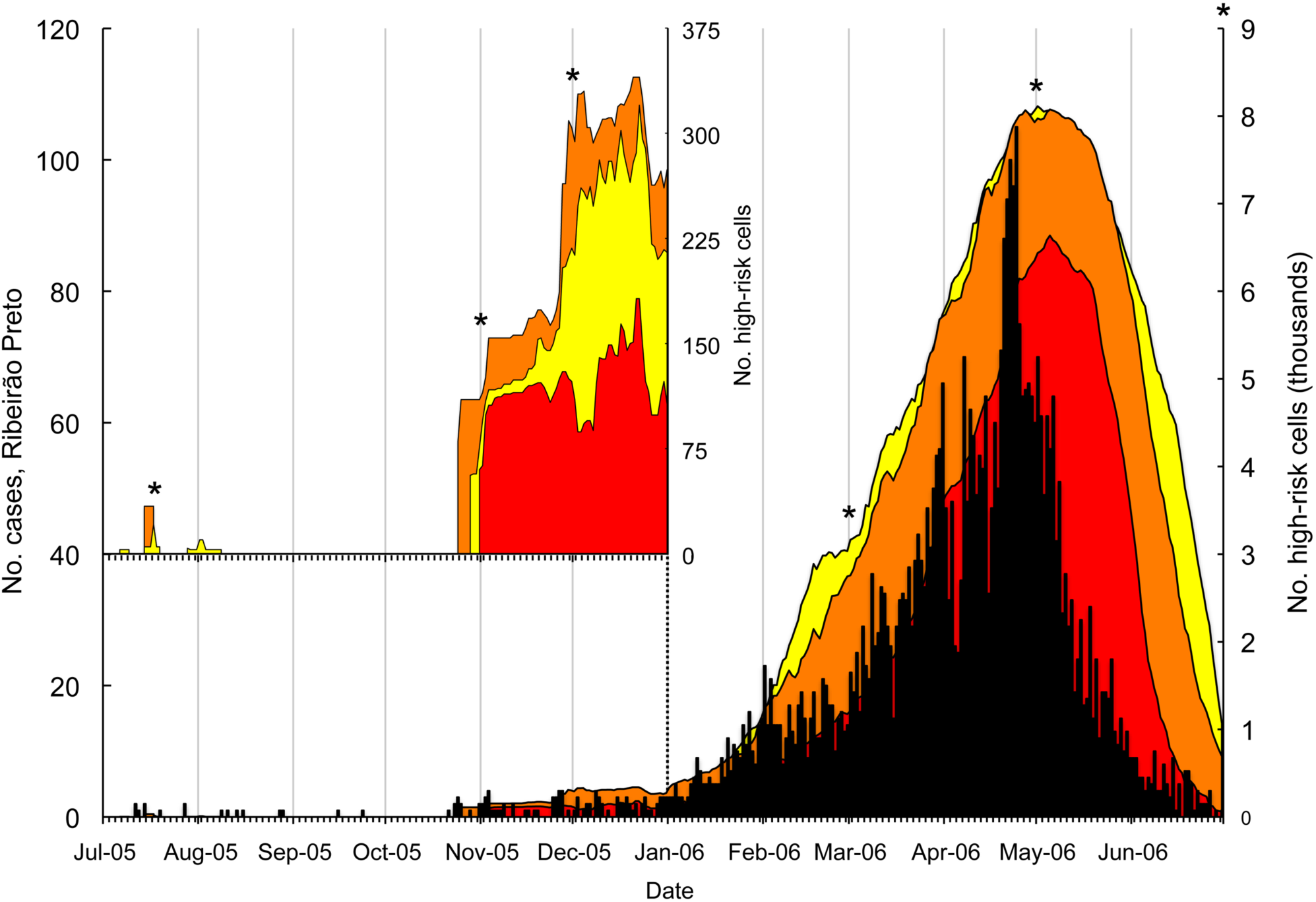
Number of confirmed dengue cases and DYCAST cells identified as high risk by models D10 (red), D05 (orange), and D10’ (yellow) per day, Ribeirão Preto, Brazil, Jul 1, 2005–Jun 30, 2006. Interepidemic period is Jul 1–Oct 31. Inset x-axis has the same scale, Jul 1, 2005–Jan 1, 2006; order of models D05 and D10’ has been reversed in the inset for visualization purposes. Asterisks denote dates at which risk maps are shown in Figure 6.

**Figure 6.**
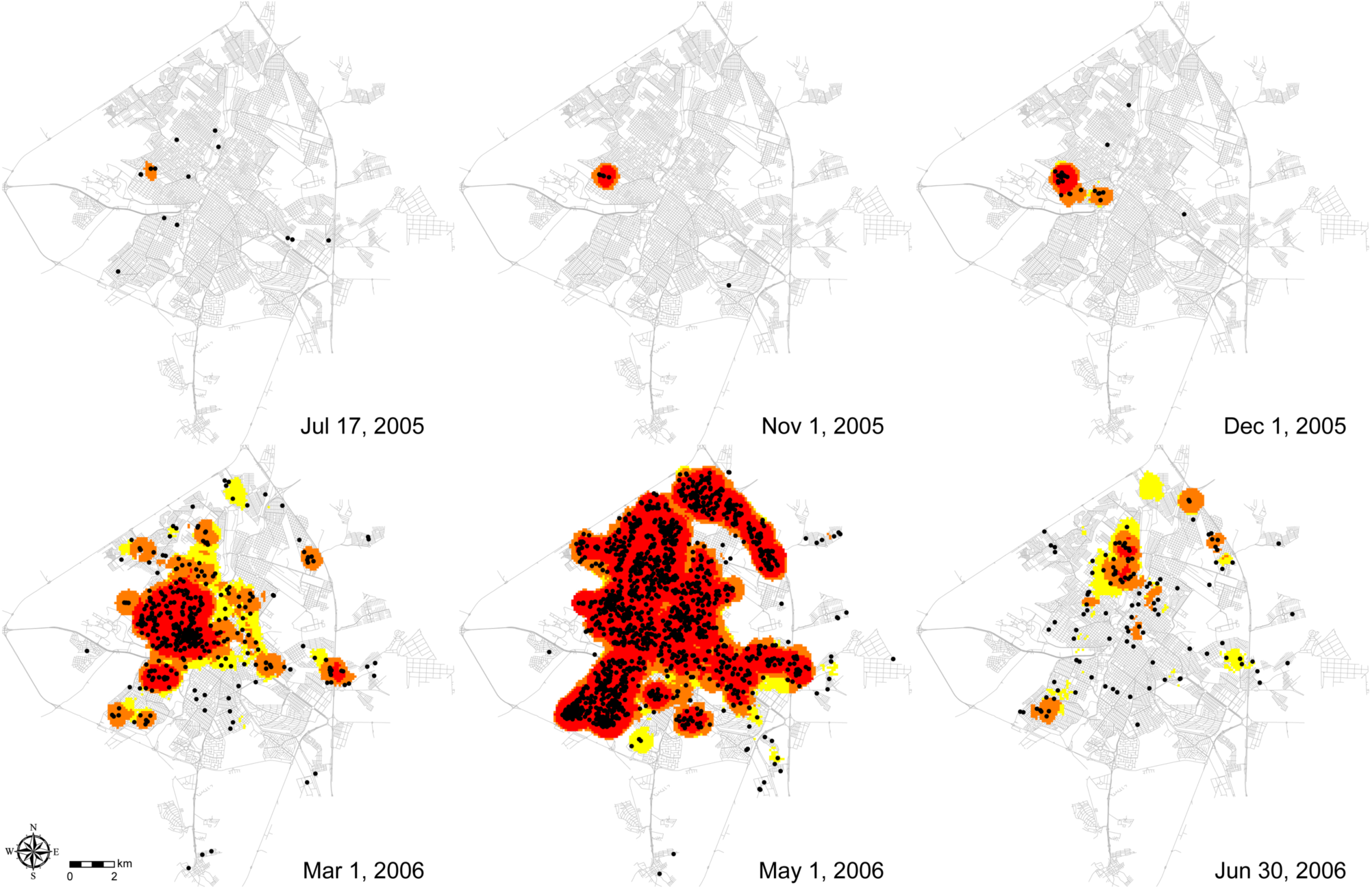
Confirmed dengue cases and DYCAST risk maps displaying high-risk cells as identified by models D10 (red), D05 (orange), and D10’ (yellow), Ribeirão Preto, Brazil. Dates correspond to asterisks in Figure 5; interepidemic period is Jul 1–Oct 31 (top left). Maps illustrate cryptic interepidemic focus of transmission (Jul 17) as well as its presumed reemergence (Nov 1) and spread to surrounding areas (Dec 1), followed by increasing (Mar 1), peak (May 1), and contracting (Jun 30) levels of transmission, as defined by the number of high-risk cells. Cases displayed are those with an onset of illness on or up to 28 days prior to given date; this corresponds to the temporal domain of the model and thus includes all cases contributing to risk at that time (see Supplementary Video for an animation of D10’ risk maps during entire study period).

## 4. Discussion

### 4.1. Summary

Given the lack of approved drug prophylaxis and specific antiviral treatment for DENV, mosquito control and personal protection measures are critical to reducing human morbidity and mortality. Thus, early warning of high-risk areas would allow such efforts to be targeted in a timely and effective manner. Results from this study indicate that the modified DYCAST model provided early and accurate identification of high risk areas in Ribeirão Preto, including detection of what appears to be a cryptic interepidemic focus of transmission that later developed into the severe 2006 epidemic. Additionally, model efficacy was retained and even enhanced by including unconfirmed dengue cases in the analysis, which has important implications for increasing the utility and applicability of the model. These findings suggest that the DYCAST system could be utilized prospectively and in real time to identify areas at high risk of DENV transmission, in order to target mosquito control, surveillance, and public education campaigns in a timely, efficient, and cost-effective manner. Furthermore, and in a departure from previous studies, this risk model was implemented using free, open-source, and cross-platform software that could provide an inexpensive and scalable GIS solution for the surveillance and control of DENV – and potentially other infectious diseases – by Ribeirão Preto and other public health agencies around the world.

### 4.2. Significance

The DYCAST models’ average sensitivity (across all specificities) of 78.3% is given by the AUC (0.783) (*42*). However, this should be considered an approximation, as only three data points were used to plot the ROC; the D10’ model used different inputs than the other models as well, albeit its inclusion resulted in only a negligible increase in AUC (0.003). Youden’s J statistic, the sum of each model’s sensitivity and specificity over that of random chance, ranges from 0 (random chance) to 1.0 (perfect agreement; also -1.0 for perfect disagreement) and is useful if sensitivity and specificity are of equal importance in determining an optimal threshold (*43*). Model D10’ had the highest J index as well as sensitivity, and would therefore be considered the optimal model; only if specificity were of primary concern would model D10 be considered optimal.

Sensitivity values and prediction rates are interpreted to be quite high, given that the dengue models use cases to predict future cases, and as such the incipient cases will inherently reduce both measures by never being predicted (albeit this may have been partially offset by considering as predicted any cells identified as high-risk on the date of onset of its earliest case). Furthermore, both the sensitivity and prediction rate of models D10’ and D05 are greater than that of the WNV DYCAST model implemented during the unprecedented 2005 epidemic in California. This model, which used reports of dead birds to predict human cases, had a sensitivity of 80.8% (269/333 cells) and a prediction rate of 81.6% (289/354 cases) (*19*). However, specificity (90.6%; 66,543/73,434) was higher than that of all three dengue models.

These differences between the dengue and WNV models may reflect dissimilar viral ecologies. As a zoonotic disease, WNV transmission may be curtailed through die-off and migration of infected and susceptible avian hosts, dynamics that do not apply to the DENV cycle. Rather, continuous presence of dense human populations, coupled with much smaller flight range and domestic proclivities of the *Ae. aegypti* vector, presumably account for more sustained viral transmission at a smaller scale. Indeed, cells were identified as high-risk for a much longer total mean duration by the dengue models (65.4–92.3 days) than by the WNV model (39.0 days) (*19*). This phenomenon may also account in part for the observation that during the study period, half of all cases (50.1%; 2,352/4,690) occurred in cells containing a prior case; 44.4% of cells (1,039/2,338) that contained at least one case contained multiple cases (mean 3.3, maximum 31; mean across all occupied cells 2.0). Of course, these findings may also be explained by the fact that a single *Ae. aegypti* mosquito can infect multiple persons within the same residence.

Regardless, this redundancy of cases within single cells accounts for the differences between prediction rates (percentage of cases predicted) and model sensitivity (percentage of cells that predicted their earliest case out of all cells that contained cases). Before rounding, prediction rates exceeded sensitivities by 12.0, 7.1 and 6.6 percentage points for models D10, D05, and D10’, respectively. Additionally, this redundancy allows for prediction rates to be compared against a simple null “model,” whereby cells are identified as high risk once a case occurs within them. This is equivalent to the aforementioned percentage of cases that occurred in cells containing a prior case, which provides a prediction baseline of 50.1% (2,352/4,690 cases). Models D10, D05, and D10’ thus had prediction rates ≈1.6–1.8 times higher than that of the null model, which equates to an additional 1,358 (29.0% of all cases), 1,882 (40.1%), and 1,885 (40.2%) cases predicted, respectively.

Results from all three models also indicated relatively high accuracy (percent agreement), as well as a moderate strength of chance-adjusted agreement (0.40<kappa<0.60, *44*); kappa can be interpreted as the percentage of correctly identified cells expected to be misidentified by chance alone. PPV and NPV are a function of the prevalence of the outcome of interest, thus the low prevalence of cells that contained a human case (9.2%; 2,338/25,487) resulted in relatively low PPV and high NPV. Because cell RR is not influenced by prevalence, it is a more useful (albeit proxy) measure in this circumstance for communicating model predictiveness. This measure indicates that the RR of containing a confirmed case was >10× higher in cells identified as high-risk compared to those not identified as such.

One of the most surprising findings in this study was that running the model with both confirmed and unconfirmed cases provided earlier and greater prediction of human cases, as well as higher sensitivity and RR, than using confirmed cases alone. These additional records represented an increase of 95.9% (4,496/4,690 records) over the confirmed cases, and consisted of seemingly useless information – 96.4% (4,335/4,496) of unconfirmed cases were classified as negative for dengue through clinical diagnoses or laboratory testing. However, the efficacy (and superiority) of model D10’ may be explained by the following two factors. First, the model’s statistical procedure for identifying significant clustering of cases in space and time should effectively eliminate the random noise in the data, and the fact that the procedure is based on dengue-specific parameters should reduce detection of significant clustering due to non-dengue illnesses. Secondly, there may actually be an epidemiologic signal of dengue in the non-confirmed case data. This may be due to the presence of undetected dengue cases within records that were missing data (n=159) or due to false clinical diagnoses or laboratory findings that were negative (n=4,335) or inconclusive (n=2) for dengue. This is supported by the fact that unconfirmed cases generally follow annual trends of dengue incidence (Supplementary Figure S5), however this may primarily be an artifact of greater vigilance and medical attention-seeking behavior by the population during dengue epidemics.

However, it is important to note that the increase in sensitivity afforded by D10’ comes at a 6.5% reduction in specificity compared to D10 (78.8-72.3%). Presumably, this is due to the contribution of non-dengue cases to the detection of artificially significant clustering events. One solution would be to run multiple models at the same time (Figure 6), which would maximize both sensitivity and specificity. Alternatively, each model could be run at different times of the year when different indices are preferred, such as running the more sensitive D10’ during the interepidemic period and running the more specific D10 during the epidemic period. Furthermore, because D05 also provides better prediction and sensitivity than D10, it may be possible that a further increase in sensitivity could be achieved by creating a hybrid of D10’ and D05 that would analyze all suspected cases at a threshold of 5.

From a practical standpoint, these results suggest that the model may retain efficacy in areas where thorough testing is not available or practical due to lack of resources or infrastructure; this has implications for making DYCAST applicable to a wider audience, as well as for the utility of modeling syndromic surveillance data. Additionally, the ability to predict high-risk areas without having to confirm suspected dengue cases eliminates the delay incurred from laboratory testing and clinical diagnoses, providing more timely results and thus a more rapid response to disease transmission.

Even without such an advantage of earlier case reporting, model D10’ predicted cases with a greater mean number of days prior to onset than did the other models. Regardless, all models predicted cases with a mean length of time prior to onset sufficient for mosquito control efforts to respond and potentially interrupt viral transmission, even accounting for the 6-day human incubation period. The models’ ability to provide early warning of disease transmission appears not to be a simple consequence of the large number of multiple cases in single cells (which increases the average duration); based only on the earliest cases within cells, the mean number of days that a cell successfully predicted its earliest case was 55.7 (std dev 38.0, median 51, max 292), 51.4 (std dev 38.5, median 46, max 295), 31.7 (std dev 31.7, median 32, max 201) for models D10’, D05, and D10, respectively.

Another important finding is that as early as July 2005, DYCAST identified what appears to be the primary interepidemic focus of transmission that preceded the severe 2006 epidemic. This suggests the possibility that high-risk areas identified during the interepidemic period may reflect overwintering of the virus. Another notable feature of this detection is that without the risk maps, the clustering of cases is not conspicuous, and in fact appears against a backdrop of other seemingly scattered cases (Figure 6, Jul 17, 2005). This indicates that the model’s statistical approach may add value to current surveillance and control efforts. Ultimately, such detection of otherwise cryptic transmission foci provides an opportunity for targeted mosquito control efforts to interrupt viral amplification before it reaches “critical mass” and spills over to surrounding areas.

### 4.3. Limitations

One limitation of the study includes potential selection bias of dengue cases, in that addresses not included in the street reference data, such as those in the rural periphery of the town or in newly developed areas, would not be geocoded. This may affect the case incidence rate as well as the sensitivity of the risk model. Future work could identify the presence of this bias through assessment of spatial structuring of geocoding rates. Additional bias could result from a patient’s working address being recorded instead of their residential address. While difficult to assess posteriorly, this and similar errors could be mitigated through quality control and manual rematching of unmatched addresses by someone local and familiar with the area.

A related bias concerns one of the primary assumptions of the model, which is that illness is acquired at a person’s place of residence. While this is particularly consistent with the domestic nature of DENV transmission, illnesses acquired outside of a person’s residence may confound the results. The second assumption of the model, that DENV transmission is spatially continuous, may be violated by virtue of the fact that persons can also transmit the virus to mosquitoes outside of their place of residence as well. Provided sufficient rates of vertical transmission, this assumption may also be violated by accidental human transport of *Ae. aegypti* eggs and larvae, which is reported in Brazil to be more important than adult flight for dispersion of the vector (*29*). Nevertheless, given the spatially confined flight range of the adult form that actually transmits the virus, violations of this assumption are considered to be negligible.

The third assumption of the model is that non-random spatiotemporal clustering indicates amplification of DENV. While this assumption is fully met by using only confirmed dengue cases, it may be violated by including non-dengue cases, as likely illustrated by the 6.5% reduction in specificity of D10’ compared to that of D10. However, as mentioned above, the model’s dengue-specific statistical procedure should generally limit the influence of such violations. Fourth, it is assumed that each dengue case has an equal opportunity of being reported. Differential rates of symptomatic DENV illness among various immunologic, age, or demographic groups may violate this assumption if such populations exhibit sufficient spatial heterogeneity (e.g., distribution of DENV antibodies within the population following spatially-structured epidemics). This and the fact that DENV is asymptomatic in the majority of infections (*45*) may also yield false positive identifications, thereby reducing validity measures such as specificity and PPV. Unfortunately, these and other reporting biases are difficult to rigorously assess in the absence of serologic surveillance information. Symptomatological considerations aside, however, the existence of free universal healthcare in Brazil and prevalent media campaigns informing citizens about DENV symptoms may ensure that this assumption is generally met.

Other limitations concern the implementation of the software itself. The tradeoff with using multiple open source software components is that compatibility issues may arise as new versions of the various components are released. Additionally, running the model is computationally expensive, and a computer with powerful hardware is recommended.

### 4.4. Future implementation

One important avenue for future research includes the biology of *Ae. aegypti* from local populations, as findings from previous studies are often inconsistent and/or location-specific. Three potential research objectives for informing the DYCAST model parameters are recommended. First, mark-release-recapture (MRR) studies could determine the mean and maximum dispersion of the vector, which would elucidate the optimal spatial domain and spatial closeness parameters for a particular study area. Second, research could examine the role of temperature in affecting the lengths of the EIP and gonotrophic cycle of *Ae. aegypti*, which would inform the temporal domain and temporal closeness parameters. Such findings could be incorporated by adjusting these parameters based on real-time temperature data, or by specifying longer and shorter temporal parameters for winter (interepidemic) and summer (epidemic) models, respectively. Third, population census data could be sampled at each grid cell’s centroid at a radius equal to the model’s 600 m spatial domain, in order to generate Monte Carlo distributions that reflect the spatial heterogeneities of the underlying human population (*46*). A more sophisticated approach, dasymetric mapping (*47*), could be used to re-aggregate population counts to only populated regions within a given census tract, as defined by remote sensing-derived land use classification imagery and/or residential lots data. This would yield a more accurate population layer by reducing error from the ecological fallacy of choropleth mapping.

Recommendations for future program development include the establishment of a pilot program to create an open source GIS network for electronic mapping and sharing of otherwise fragmented datasets used for surveillance and control by the various local health departments, vector control agency, and municipal government (CODERP) in Ribeirão Preto. This would provide for routine and timely mapping of surveillance data, and would allow for more rapid, coordinated, and efficient management across all facets of disease control, from public education campaigns and clinical surveillance to prospective modeling and targeted vector control. This integrated framework would also better facilitate the practical application of the current research findings and act as a catalyst for further in-depth research.

This program would also serve as a replicable template for deploying scalable, no-cost GIS solutions that can be adopted by resource-limited areas around the world for the surveillance and control of dengue and other infectious diseases. Rather than relying on expensive commercial software packages, this project could leverage the free open source software solutions used in the present study, thus providing an inexpensive but powerful GIS network with potential for web and mobile deployment. Future surveillance data could be collected by local agencies and entered in electronic format via PostgreSQL clients or online interfaces, created by using the Python Web Processing Service (PyWPS), for access remotely or via mobile devices; dynamic web content, such as case and risk maps used for public education campaigns, could be published by using Hypertext Preprocessor (PHP) scripting language. Security could be maintained by using Secure Sockets Layer (SSL) protocol to encrypt client/server communications, as well as password protection of computer accounts. Case data entered from the backend could be automatically geocoded and mapped; support for Geospatial Data Abstraction Library (GDAL) via the GDAL-GRASS plug-in would enable integration with other software as needed. Quantum GIS (QGIS; www.qgis.org) could be implemented for a more user-friendly frontend that includes a GUI in Portuguese, or GRASS GIS, which supports various visualizations (2D, 2.5D, and 3D raster and vector data) and spatial analyses. The latter could include R (www.r-project.org), coupled with the crimestats, spatstat, and splancs packages for additional spatial and spatiotemporal analyses.

Regardless of these additional features, installation of the components used in this study would allow for the prospective implementation of the dengue DYCAST model, which could generate daily risk maps in real time for the immediate use in surveillance and control efforts. To facilitate global dissemination of the model and results, open-source versions of the DYCAST software are available at www.DYCAST.org, via GitHub repositories of version 2.0 (Python/PostGIS; https://github.com/almccon/DYCAST) adapted herein, and version 3.0 (Python/PostGIS, with Docker support and a QGIS plugin; https://github.com/CarneyLab). Ultimately, the practical application of these modeling efforts may act as a catalyst for further research and open-source collaboration, and enhance public health infrastructure of the developing world that bears the greatest burden of dengue morbidity and mortality.

## Supporting information

Supplementary Video. DYCAST dengue risk map animation. DYCAST high-risk cells (red) and dengue cases (black) in Ribeirao Preto, Brazil from 2005-07-01

## Data Availability

All data produced are available online at

https://www.DYCAST.org

## Acknowledgements

Funding for this research was provided through a U.S. Centers for Disease Control and Prevention Training Grant Master of Public Health Fellowship, with subsequent development of DYCAST version 3.0 supported by National Science Foundation grant number IIS-2014547. This manuscript is derived from my MPH thesis completed at Yale University. I would particularly like to thank my academic and thesis advisor, Durland Fish, PhD, for his valuable direction and insight, as well as preceptor Benedito Antônio Lopes da Fonseca, MD, PhD, MPH (Universidade de São Paulo) and colleague André Markon, MPH (University of Michigan) for their exceptional assistance during my summer internship. I would like to thank my thesis reader Maria Diuk-Wasser, PhD and professor Ted Holford, PhD (Yale University) for helpful comments and feedback, and acknowledge the authors of the open-source DYCAST scripts, Alan McConchie, MS (version 2.0; University of British Columbia) and Vincent Meijer, MA (version 3.0; University of South Florida). For their assistance in providing data and information used in this study, I would like to thank Marcelo Raspa, Claudio Souza de Paula, MD, and Maria Luiza, MD (Secretaria Municipal de Saúde); Antônio Pazin Filho, MD (Universidade de São Paulo); and Wilson Yong (Companhia de Desenvolvimento Econômico de Ribeirão Preto).

## Supplementary Material

### Supplementary Video

#### DYCAST dengue risk map animation

DYCAST high-risk cells (red) and dengue cases (black) in Ribeirão Preto, Brazil from 2005-07-01 to 2006-06-30 (model D10’)

**Supplementary Table S1.** Number of confirmed dengue cases, Ribeirão Preto.

| Year | Total cases | Classic dengue | Dengue with complications | DHF | DSS |
|---|---|---|---|---|---|
| **2009\*** | **1,685** | **1,671** | **5** | **9** | **0** |
| **2008** | **1,073** | **1,066** | **3** | **2** | **2** |
| **2007\*** | **2,752** | **2,733** | **15** | **4** | **0** |
| **2006\*** | **6,051** | **6,026** | **11** | **13** | **1** |
| 2005 | 645 | 639 | 1 | 5 | 0 |
| 2004 | 50 | 50 | 0 | 0 | 0 |
| **2003** | **808** | **800** | **0** | **8** | **0** |
| 2002 | 360 | 360 | 0 | 0 | 0 |
| **2001\*** | **3,217** | **3,216** | **0** | **1** | **0** |
| 2000 | 211 | 211 | 0 | 0 | 0 |
| 1999 | 320 | 320 | 0 | 0 | 0 |
| 1998 | 268 | 268 | 0 | 0 | 0 |
| Total | 17,440 | 17,360 | 35 | 42 | 3 |

**Boldface** denotes epidemic years (https://www.ribeiraopreto.sp.gov.br/files/ssaude/pdf/pms-rp-2014-2017.pdf)
\* denotes years of critical transmission, defined by the Brazilian Ministry of Health as incidence rates >300 per 100,000 (de Paula, pers. comm.)

**Supplementary Table S2.** Sensitivity analysis of geocoding parameters using the composite address locator.

| Parameters* | Matched+Tied | Matched | Tied | Unmatched |
|---|---|---|---|---|
| 80-10-60† | 57% | 30,006 (56%) | 589 (1%) | 22,661 (43%) |
| 80-10-60‡ | 14% | 7,488 (14%) | 124 (0%) | 45,644 (86%) |
| 80-10-60 | 58% | 30,503 (57%) | 598 (1%) | 22,155 (42%) |
| 40-10-60 | 59% | 30,958 (58%) | 606 (1%) | 21,692 (41%) |
| 40-10-30 | 69% | 35,866 (67%) | 777 (1%) | 16,613 (31%) |
| 80-10-23 | 69% | 35,732 (67%) | 903 (2%) | 16,621 (31%) |
| **60-10-23** | **72%** | **37,544 (70%)** | **940 (2%)** | **14,772 (28%)** |
| 50-10-23 | 74% | 38,485 (72%) | 1,071 (2%) | 13,700 (26%) |
| 40-10-23 | 76% | 39,114 (73%) | 1,558 (3%) | 12,584 (24%) |
| 20-10-23 | 81% | 39,474 (74%) | 3,529 (7%) | 10,253 (19%) |
| 40-10-15 | 77% | 39,420 (74%) | 1,573 (3%) | 12,263 (23%) |
| 20-10-15 | 81% | 39,800 (75%) | 3,551 (7%) | 9,905 (19%) |
| 20-5-15 | 81% | 39,800 (75%) | 3,551 (7%) | 9,905 (19%) |
| 20-5-5 | 82% | 40,073 (75%) | 3,568 (7%) | 9,615 (18%) |
| 5-5-5 | 84% | 39,893 (75%) | 4,618 (9%) | 8,745 (16%) |
| 0-0-0 | 88% | 40,352 (76%) | 6,490 (12%) | 6,414 (12%) |

\*Spelling sensitivity, Minimum candidate score, and Minimum match score, respectively. Default values are 80-10-60. **Boldface** denotes optimal parameters; the final set was further modified to 60-10-10 (see text)
†CEP (zip code)-based address locator (individual)
‡Bairro (district)-based address locator (individual)

**Supplementary Figure S1.**
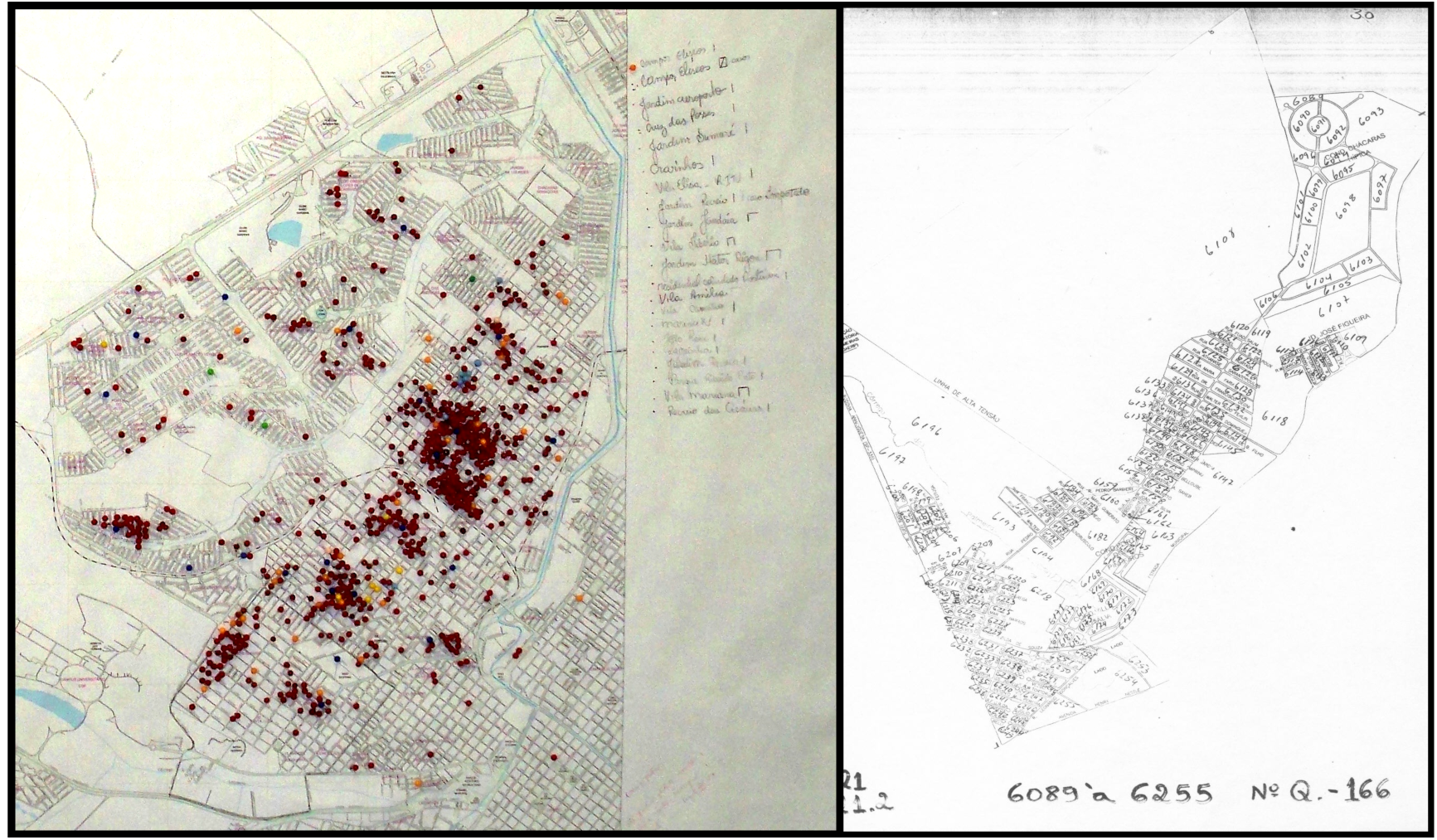
Mapping methods used in Ribeirão Preto for dengue surveillance and control in 2007. a. color-coded push pins used to plot dengue cases in one of the health districts, and b. one of dozens of papers comprising a map of the city used to track and plan for mosquito surveillance and control.

**Supplementary Figure S2.**
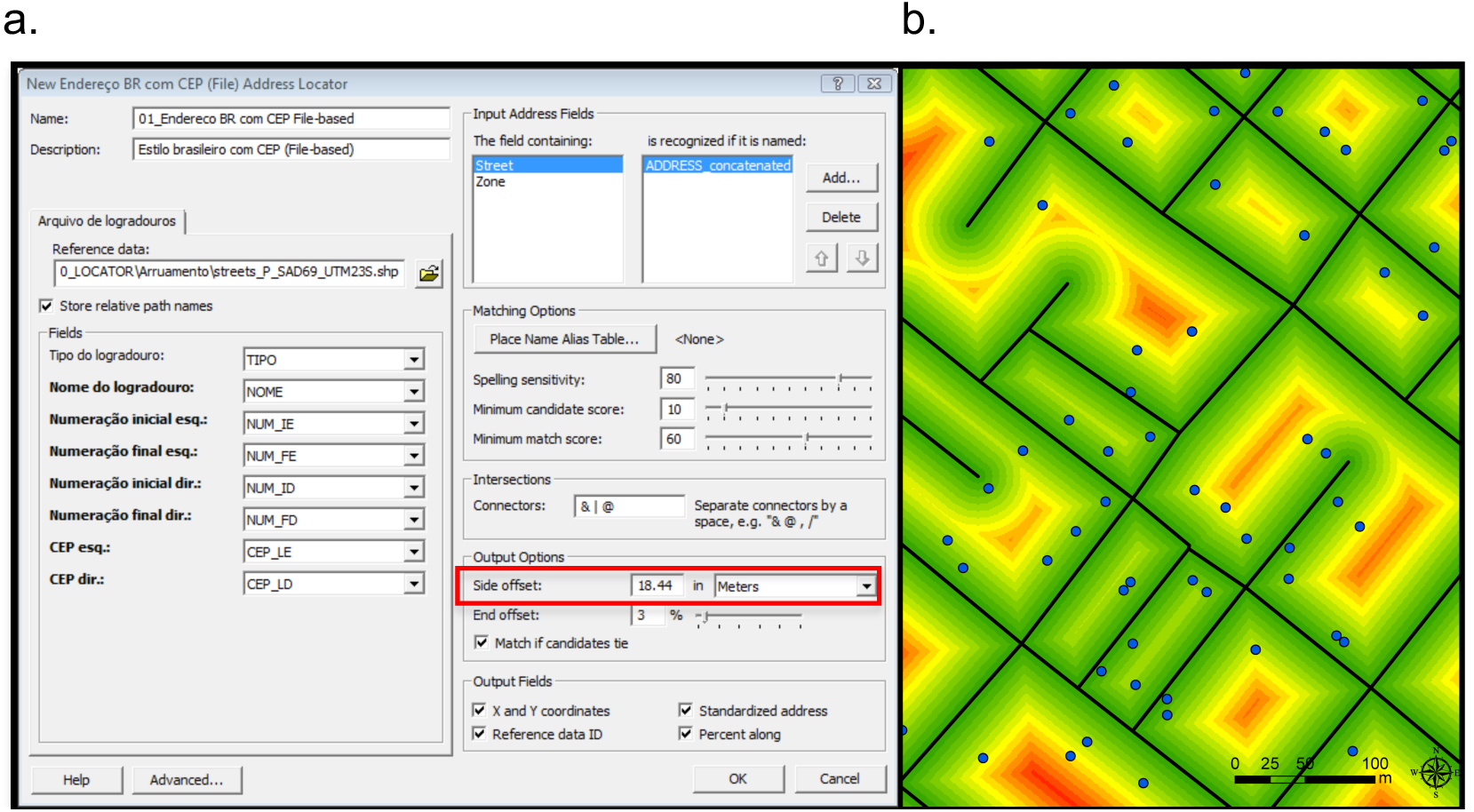
Geocoding process in ArcView 9.3.0. a. Código de Endereçamento Postal (CEP; zip code)-based address locator, illustrating matching parameters (“Matching Options”) and side offset (red box); b. raster layer of distances from street centerline (from green to red) sampled by cases (blue circles) to calculate median offset.

**Supplementary Figure S3.**
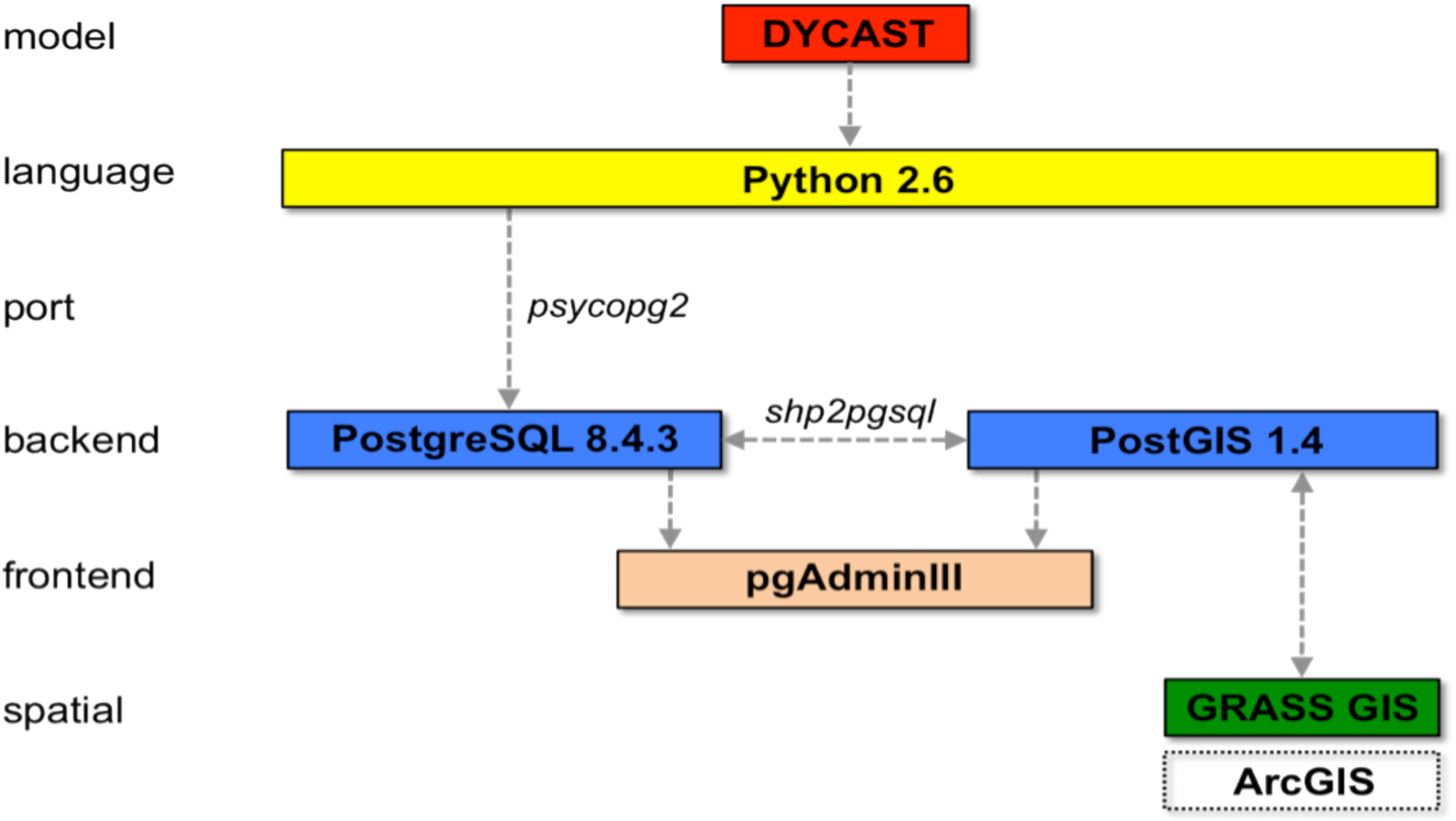
Schematic of free and open source software used to implement the DYCAST model. Commercial ArcGIS software (i.e., ArcView) was used in this study to create spatial datasets; however, this could be accomplished in future implementations by using free and open source GIS software, GRASS GIS.

**Supplementary Figure S4.**
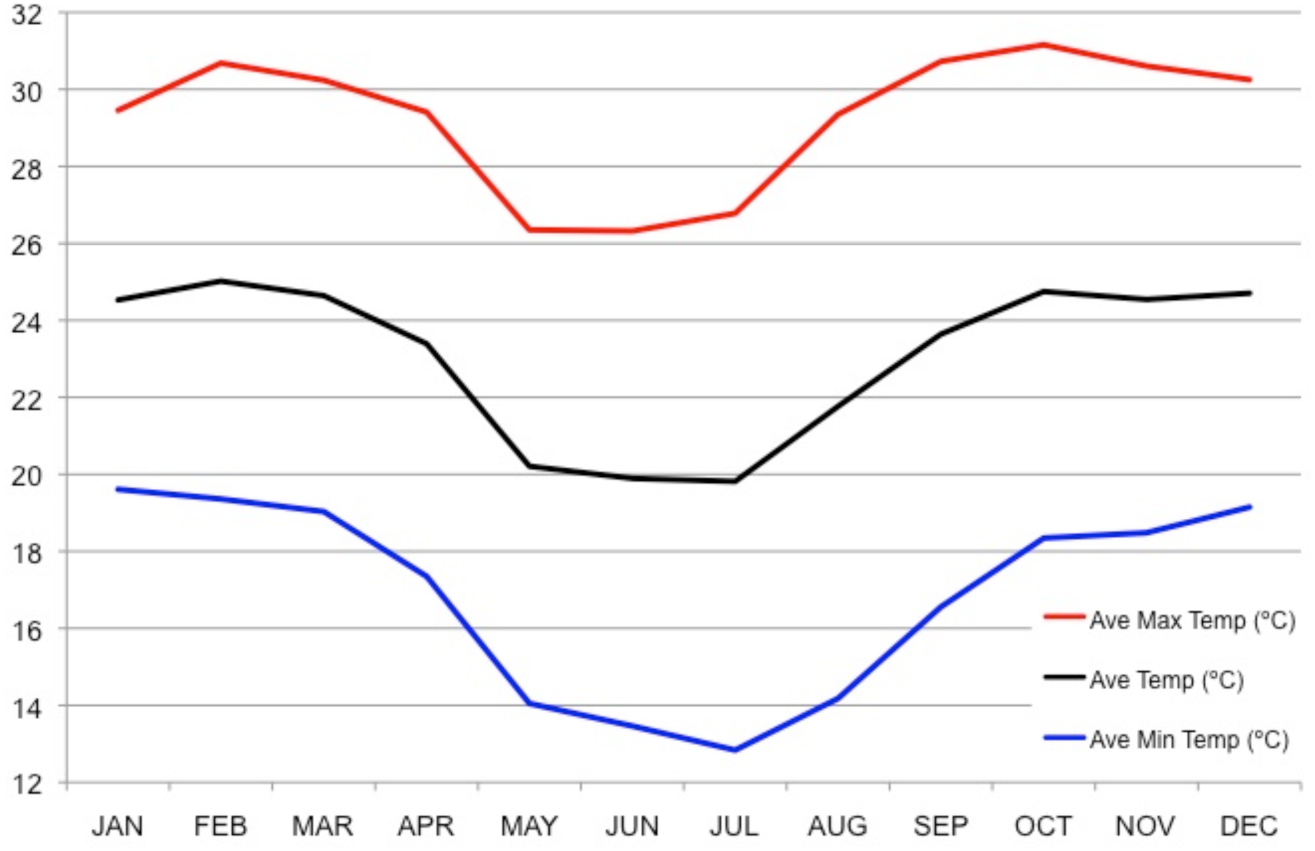
Average monthly temperatures, Ribeirão Preto, 2003-2009. Data source: Centro Integrado de Informações Agrometeorológicas (http://www.ciiagro.sp.gov.br/ciiagroonline/Listagens/MonClim/LMClimLocal.asp)

**Supplementary Figure S5.**
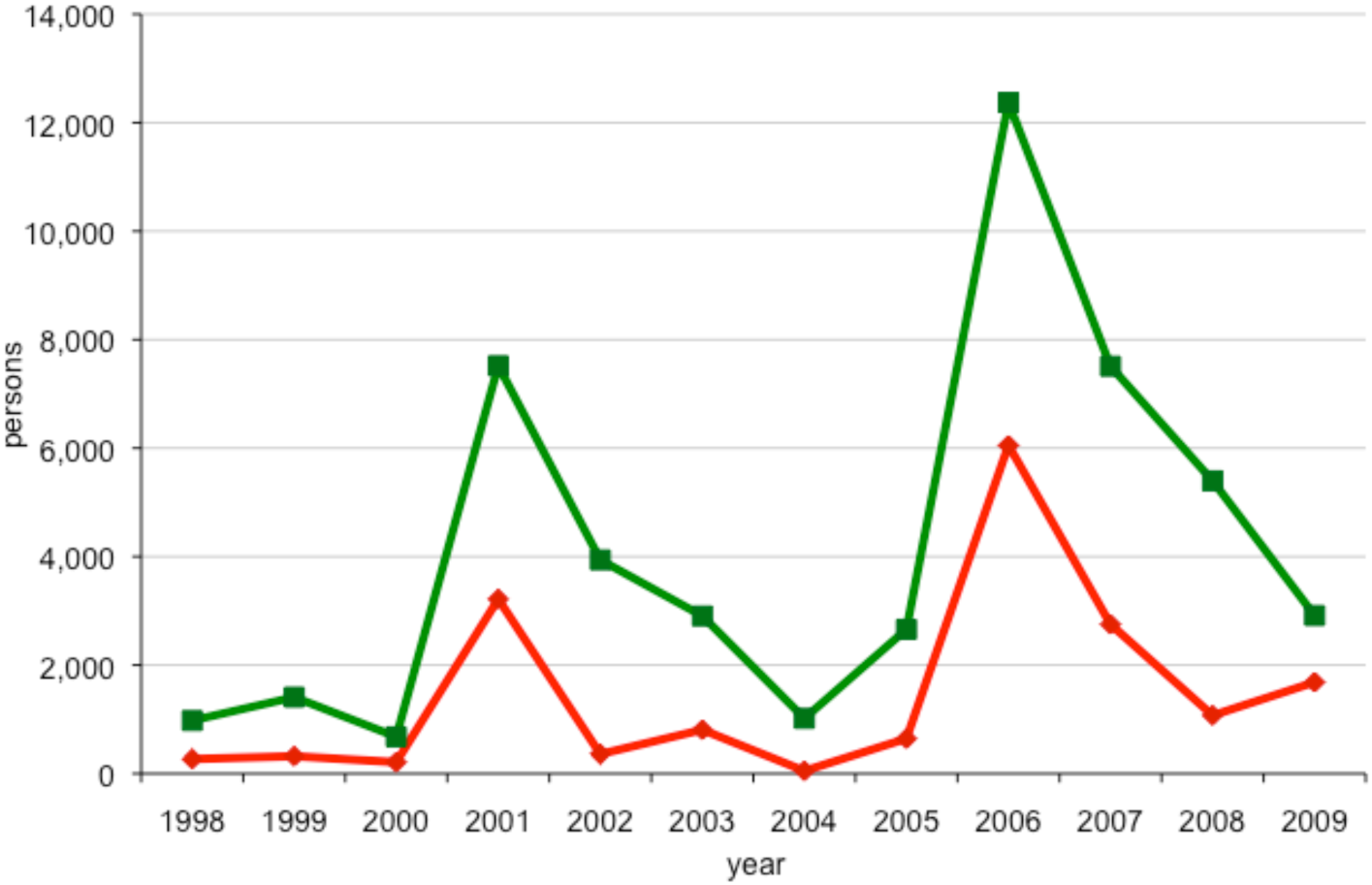
Number of suspected cases confirmed (red) and unconfirmed (green) for dengue infection per year, 1998-2009, Ribeirão Preto.

## Notes

### Competing Interest Statement

The authors have declared no competing interest.

### Author Declarations

University of South Florida IRB approval #Pro00031029: "Mosquito-borne disease early warning system and spatiotemporal analyses"

